# Frequency of Hematologic and Solid Malignancies in the Family History of Patients with Myeloid Neoplasms

**DOI:** 10.1101/2022.01.15.22269343

**Authors:** Alaa Ali, Batool Yassin, Ali Almothaffar

## Abstract

**Background:** Studies demonstrated that there are several germline mutations that lead to a familial predisposition for acute myeloid leukemia and Myelodysplastic syndrome. According to the American Society of Clinical Oncology ,the minimum cancer family history was defined as including first- and second-degree family history, type of primary cancer, and age at diagnosis.

The current study aimed to estimate the frequency of positive family history for hematologic and solid malignancies in patients with Myeloid Neoplasms / Aplastic anemia.

**Patients and Methods:** A cross section study was carried out at the Center of Blood Diseases, Medical City Campus during the period from March-December 2020. A purposeful sample of all adult patients with Myeloid Neoplasms [Acute Myeloid Leukemia, Myelodysplastic Syndrome, Chronic Myeloid Leukemia and Aplastic Anemia] were included in the study. A data collection form was prepared, based on the Hereditary Hematopoietic Malignancies Screening form adopted by the University of Chicago, and modified by the researchers; The data were collected by direct interview with the patients. Patients with hematologic malignancy and one or more first-degree relatives, or ≥2 second-degree relatives, with hematologic malignancies and individuals with Myelodysplastic Syndrome or Acute Myeloid Leukemia and two first or second-degree relatives with a diagnosis of solid tumor malignancy were considered potential carriers of such genetic predisposition.

**Results:** A total of 153 patients were included; males were nearly equal to females with a male to female ratio of nearly 1:1. Acute Myeloid Leukemia was found in 57.5%, Aplastic Anemia was found in 19%, Chronic Myeloid Leukemia in 17% and only four patients (6.5%) were known cases of Myelodysplastic Syndrome. Nine patients (5.9%) reported family history of hematological malignancies, 29 (19.0%) reported family history of solid malignancies and only one patient reported family history of both hematological and solid malignancies.

Regarding the official medical reports of the patients, no patient had been interviewed properly about this crucial point.

**Conclusion:** Positive family history for hematological and solid malignancies in Iraqi patients with myeloid neoplasms is prevalent. Our current approach to this critical issue in Iraq needs to be re-considered.

## Introduction

Studies demonstrated that there are several germline mutations that lead to a familial predisposition for acute myeloid leukemia (AML) and Myelodysplastic syndrome (MDS) ^(1-3)^. These findings have led to the implementation of a new category in the revised World Health Organization (WHO) classification of 2016 for myeloid neoplasms and acute leukemia namely myeloid neoplasms with germline predisposition as well as in the recent European Leukemia Network (ELN) classification named familial myeloid neoplasms ^(4,5)^. Nevertheless, familial cases of AML and MDS are only seldom encountered in the clinical routine and the question remains if there are a relevant number of unidentified cases as the family histories were not taken elaborately enough. The family history is an integral part of the patient history of cancer patients. The American Society of Clinical Oncology recommends as minimum family history for individuals with cancer to obtain information about first and second-degree relatives both on maternal and paternal sides, ethnicity, age at cancer diagnosis and type of primary cancer for each cancer case in the family. Moreover, the minimum adequate cancer family history should encompass results of any cancer predisposition testing in any relative. This information should be taken at diagnosis and updated in the follow-up ^(6)^.

The aim of this study was to estimate the frequency of positive family history for hematologic and solid malignancies in adult Iraqi patients with Myeloid Neoplasms / Aplastic anemia.

### Patients and Method

A cross section study was carried out at the Center of Blood Diseases, Baghdad Teaching Hospital, Medical City Campus during the period from March-December 2020 for 153 patients with history of (AML, CML,MDS and Aplastic anemia).

A data collection form was prepared, based on the Hereditary Hematopoietic Malignancies Screening form adopted by the University of Chicago ^(7)^, and modified by the researcher and supervisors. The data was collected by direct interview with the patients. A review of patients’ records was done to confirm the diagnosis and to search for written information about family history in the records. The data collection form consisted of two parts:

**Part 1:** Demographic and social characteristics of the patients and their diagnosis includes:

Patients’ ID, gender, age in years, marital status (single, married, widow, or divorced), number of children, number of siblings, diagnosis and age at diagnosis: **Part 2:** Screening questions for the patients and his / her first degree and second-degree relatives which includes:

Hematological and solid malignancies, Chronic blood diseases, blood transfusion, bone marrow failure, lymphedema, serious infections necessitate hospital admissions, warts, neurological and skeletal abnormalities. A copy of the form is available on request.

Patients with a positive family history of haematological or solid malignancies or both were sorted for the possibility of familial germline predisposition to myeloid neoplasms, among them those with one or more of the following criteria were considered to be suspected cases that should have been referred for genetic studies^(8)^:

❖ A hematologic malignancy patient with one or more first-degree relatives, or ≥2 second-degree relatives, with hematologic malignancies.
❖ One individual with MDS or AML and two FDR or SDR with a diagnosis of solid tumor malignancy.

### Statistical analysis

The Data was summarized, presented and analyzed using Statistical Package for Social Sciences (SPSS) version 21 and Microsoft Excel, 2016. Continuous variables were presented as mean ± standard deviation, and categorical variables were presented as frequency and relative frequency. Chi square test was used to test the significant association between categorical variables, student’s t test and Analysis of Variance (ANOVA) were used to test the significant differences between means. P value of 0.05 was considered statistically significant.

## Results

In the current study 153 patients were included; males were nearly equal to females with a male to female ratio of nearly 1:1, 76.5% were ever married; among them 74.4% with three children or more and only four patients were with no siblings (table1).

**Table (1):** Basic demographic data of the participants.

| **Characteristics** | **No.** | **%** |  |
| --- | --- | --- | --- |
| **Gender** |  |  |  |
| **Male** | 76 | 49.7 |  |
| **Female** | 77 | 50.3 |  |
| **Total** | 153 | 100.0 |  |
| **Social status** |  |  |  |
| **Single** | 36 | 23.5 |  |
| **Ever married** | 117 | 76.5 |  |
| **Total** | 153 | 100.0 |  |
| **No. of children** |  |  |  |
| **None** | 9 | 7.7 |  |
| **1-2** | 21 | 17.9 |  |
| **3-4** | 33 | 28.2 |  |
| **5-6** | 27 | 23.1 |  |
| **≥7** | 27 | 23.1 |  |
| **Total\*** | 117 | 100.0 |  |
| **No. of siblings** |  |  |  |
| **None** | 4 | 2.6 |  |
| **1-2** | 14 | 9.1 |  |
| **3-4** | 39 | 25.5 |  |
| **5-6** | 35 | 22.9 |  |
| **≥7** | 61 | 39.9 |  |
| **Total** | 153 | 100.0 |  |
| **Age in years** | **Male** | **Female** | **Total** |
| **Range** | 14-90 | 14-79 | 14-90 |
| **Mean ± SD** | 42.4 ± 19.6 | 42 ± 16.9 | 42.2 ± 18.2 |
| **Median** | 42 | 41 | 41 |

\* Excluding single participants

Mean age of males was slightly higher than females and the differences in mean age between males and females was statistically not significant (table 1).

Acute Myeloid Leukemia (AML) was found in 57.5%, Aplastic Anemia (AA) was found in 19%, Chronic Myeloid Leukemia (CML) in 17% and only four patients (6.5%) were known cases of Myeloid Dysplastic Syndrome (MDS). Although AA, CML and MDS were more among females yet the association between gender and type of the disease was statistically not significant (ꭙ^2^= 2.05, df= 3. P=0.56) (table 2).

**Table (2):** Distribution of participants by diagnosis and gender*.

| Diagnosis | Males (76) |  | Females (77) |  | Total (153) |  |
| --- | --- | --- | --- | --- | --- | --- |
|  | No. | % | No. | % | No. | % |
| Acute Myeloid Leukemia (AML) | 48 | 54.5 | 40 | 45.5 | 88 | 57.5 |
| Aplastic Anemia (AA) | 13 | 44.8 | 16 | 55.2 | 29 | 19.0 |
| Chronic Myeloid Leukemia (CML) | 11 | 42.3 | 15 | 57.7 | 26 | 17.0 |
| Myeloid Dysplastic Syndrome (MDS) | 4 | 40.0 | 6 | 60.0 | 10 | 6.5 |
| Total | 76 | 49.7 | 77 | 50.3 | 153 | 100.0 |

\*The association was statistically not significant ( $\chi^2= 2.05$ , df= 3. P=0.56)

The current study revealed that among the 29 patients with aplastic anemia only three (10.3%) reported family history of malignancy; all among second degree relatives, one (3.4%) hematological malignancy and the other two (6.9%) with solid malignancies, whereas among the 88 patients with Acute Myeloid Leukemia 27 (30.7%) reported family history of malignancy; six (6.8%) hematological malignancies (three of them were first degree relatives, two were second degree and two patients reported hematological malignancies in both 1^st^ and 2^nd^ degree relatives), 20 patients reported family history of solid malignancies (seven in 1^st^ degree relatives, 12 in 2^nd^ degree relatives and one patient reported hematological malignancies in both 1^st^ and 2^nd^ degree relatives), and only one patient reported family history of both solid and hematological malignancies in both 1^st^ and 2^nd^ degree relatives.

Among the 26 patients with Chronic Myeloid Leukemia seven (26.9%) reported family history of malignancy; two (7.7%) hematological malignancies (one in each 1^st^ and 2^nd^ degree relatives), five patients reported family history of solid malignancies (three in 1^st^ degree relatives and two in 2^nd^ degree relatives) ,as for the ten patients with Myelodysplasia only two (20.0%) reported solid malignancies in 2nd degree relatives.

In summary; table 3 showed that, among all participants, nine patients (5.9%) reported family history of hematological malignancies, 29 (19.0%) reported family history of solid malignancies and only one patient reported family history of both hematological and solid malignancies

**Table (3):**
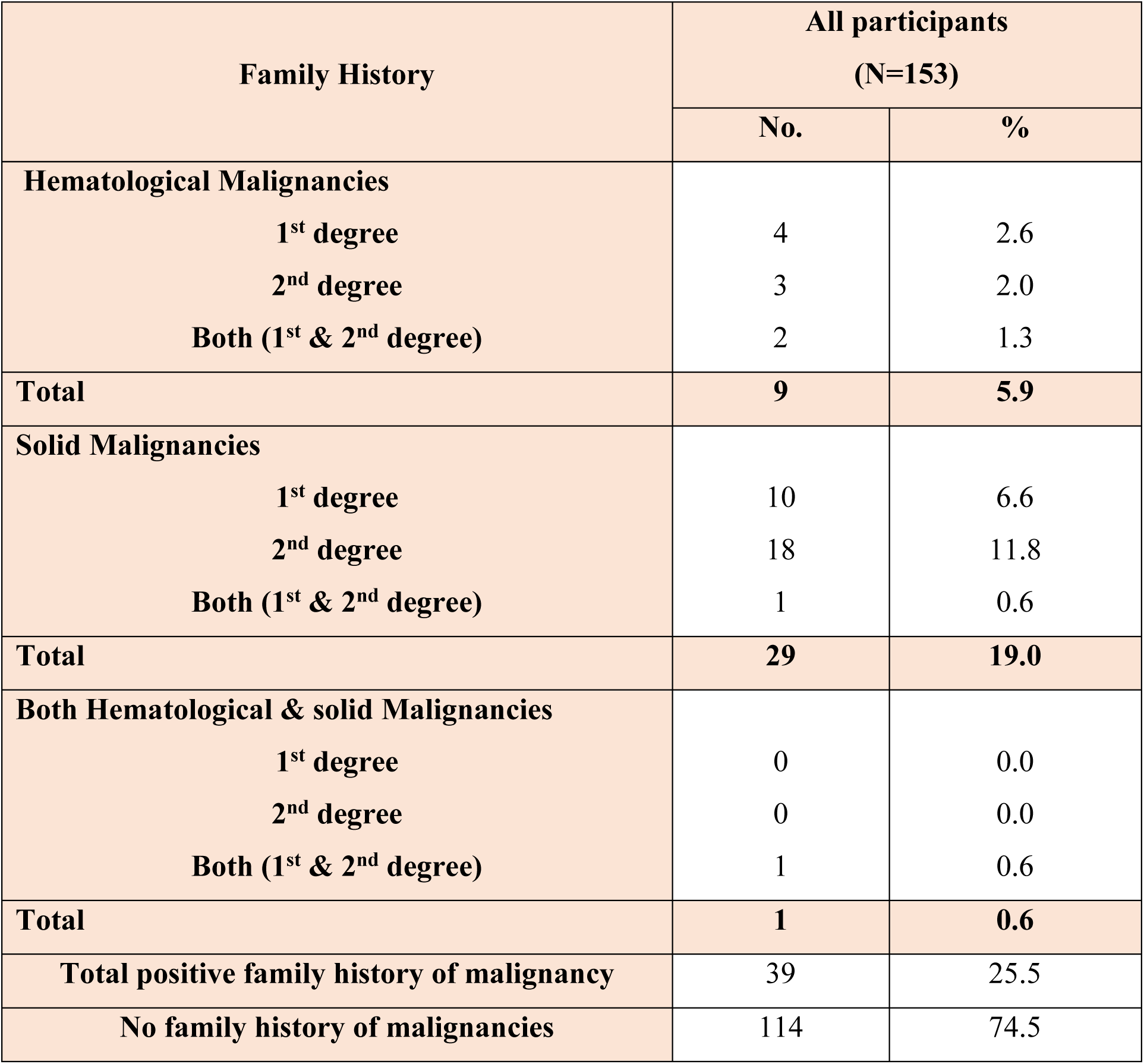
Distribution of positive family history in all the studied patients.

| Family History | All participants<br>(N=153) |  |
| --- | --- | --- |
|  | No. | % |
| **Hematological Malignancies** |  |  |
| 1<sup>st</sup> degree | 4 | 2.6 |
| 2<sup>nd</sup> degree | 3 | 2.0 |
| Both (1<sup>st</sup> & 2<sup>nd</sup> degree) | 2 | 1.3 |
| **Total** | **9** | **5.9** |
| **Solid Malignancies** |  |  |
| 1<sup>st</sup> degree | 10 | 6.6 |
| 2<sup>nd</sup> degree | 18 | 11.8 |
| Both (1<sup>st</sup> & 2<sup>nd</sup> degree) | 1 | 0.6 |
| **Total** | **29** | **19.0** |
| **Both Hematological & solid Malignancies** |  |  |
| 1<sup>st</sup> degree | 0 | 0.0 |
| 2<sup>nd</sup> degree | 0 | 0.0 |
| Both (1<sup>st</sup> & 2<sup>nd</sup> degree) | 1 | 0.6 |
| **Total** | **1** | **0.6** |
| **Total positive family history of malignancy** | **39** | **25.5** |
| **No family history of malignancies** | **114** | **74.5** |

Sorting the patients according to their family history of malignancies revealed that among those with Myeloid Neoplasms (AML, CML and MDS) eleven were suspected to have familial germline predisposition to myeloid neoplasms compared to none among patients with aplastic anemia (table 4).

**Table (4):** Patients with myeloid neoplasm or aplastic anemia who are suspected to have familial germline predisposition to myeloid neoplasms.

| Diagnosis | One or more 1<sup>st</sup> degree, two or more 2<sup>nd</sup> degree relative with |  | Two or more 1<sup>st</sup> degree, or two or more 2<sup>nd</sup> degree relative with solid malignancies |
| --- | --- | --- | --- |
|  | Hematological Malignancies | Hematological & solid Malignancies |  |
| **Patients with Myeloid Neoplasms (AML-CML-MDS)** | 4 | 1 | 6 |
| **Patients with Aplastic Anemia** | 0 | 0 | 0 |

Three patients with AML got another malignancy; the first was female got breast cancer, the second was male got lymphoma, and the third was male got thyroid cancer, yet none of them gave family history of malignancies so they were not included with above eleven patients.

Nearly the mean age (in years) of those patients with positive family history of malignancies was nearly equal to those without family history and the difference was statistically not significant (T test, df=151, P=0.822) (table 5), as for the gender table 5 showed that the association between gender and family history of malignancies was statistically not significant (ꭙ^2^= 0.05, df= 1. P=0.8).

**Table (5):** Gender and Age distribution of patients by their family history of malignancies.

| Gender | Patients with positive FH (N=39) |  | Patients with negative FH (N=114) |  | Total (153) (N=153) |  | P Value |
| --- | --- | --- | --- | --- | --- | --- | --- |
|  | No. | % | No. | % | No. | % |  |
| Males | 20 | 26.3 | 56 | 73.7 | 76 | 49.7 | 0.8* |
| Females | 19 | 24.7 | 58 | 75.3 | 77 | 50.3 |  |
| Total | 39 | 25.5 | 114 | 74.5 | 153 | 100.0 |  |
| Age in Years |  |  |  |  |  |  | 0.822** |
| Range | 14-90 |  | 14-90 |  |  |  |  |
| Mean ± SD | 40.3 ± 18.1 |  | 40.9 ± 17.1 |  |  |  |  |
| Median | 38.8 |  | 38 |  |  |  |  |

\*The difference in mean age was statistically not significant (T test, df=151, P=0.822), \*\*The association was statistically not significant ( $\chi^2=0.05$ , df= 1. P=0.8)

Reviewing Patients’ records revealed no data about family history of malignancies.

## Discussion

The current study aims to estimate the frequency of positive family history for hematologic and solid malignancies in patients with Myeloid Neoplasms / Aplastic anemia.

Nearly two thirds of the studied patients (62.8%) were with more than five siblings and 46.2% had 5-7 children, which is high compared to families in western countries and reflect the relatively large family size in Iraq that gave us more detailed family history and more chance to get information about the family of the patient and chance of getting donors for future treatment plan for the patients that includes allogeneic bone marrow transplantation.

This study showed that age of the patients and the diagnosis were nearly the same in both genders (P>0.05), same results were found by McMullin et al. 2015, on studying patient perspectives of a diagnosis of myeloproliferative neoplasm ^(9)^and Wang and Liu, 2019 on studying the pathogenesis of aplastic anemia ^(10)^

Individuals with germ line predisposition to hematologic malignancies are diagnosed with increasing frequency, the need for clinical surveillance has become apparent since presentations and disease progression can be subtle, and treatment strategies must be tailored. increasing numbers of patients and families were recognized as having an inherited cancer predisposition syndrome. Although, the first member of the family identified has already developed a hematologic malignancy, testing of additional family members results in the identification of those who have the deleterious mutation but who have not yet been diagnosed with cancer.^(11)^

In the current study we identified nine patients (5.9%) with myeloid neoplasm and aplastic anemia with family history of hematological malignancy in their first- and second-degree relatives, 29 patients (19%) with family history of solid malignancy in their first- and second-degree relatives and only one patient (0.6%) reported family history of both hematological and solid malignancy in his first- and second-degree relatives.

Zurbriggen et al., 2020, on studying hereditary hematologic malignancy syndromes in Adults, found that approximately 10 to 12 percent of adults with myeloid malignancies reported at least one close relative with a hematologic malignancy^(12)^,this was higher than what we found in the current study as only 6.5% reported family history of at least one close relative with a hematologic malignancy, also till now we lack the genetic testing required to diagnose the family members, who are healthy but harboring the genetic defect.

Sandner et al., 2019, on studying the frequency of hematologic and solid malignancies in the family history of 50 patients with acute myeloid leukemia found that 16% patients with AML had family members with hematologic malignancies, 4% of patients had family members of first degree with hematologic malignancies, in 84% of patients with AML solid malignancies were documented in family members of any degree and in 62% solid malignancy was reported in first degree relative.^(13)^ These findings were much higher than what we found in the current study as only 8% of our AML patients reported family members with hematologic malignancies, yet 6.8% of them were 1^st^ degree relative which was higher than what was found by Sandner et al. As for solid malignancies 23.9% reported solid malignancies in family members of any degree and only in 10.2% solid malignancy was reported in first degree relative. These discrepancies may indicate that family history in such patients was not considered important by both patients and physicians in these cases. In advanced international cancer centers, the family history is mapped thoroughly through extensive check list and we used a modified version of the check list from the university of Chicago to explore the family history of our patients. Surprisingly no mention about these data were found in the patients’ hospital or personal records.

Churpek et al. 2019, in their review article suggested that patients with AML should be referred for comprehensive cancer risk assessment if the following criteria apply; A family history of MDS/AML or aplastic anemia ,early onset cancer of any type or Multiple close relatives with cancer and/or a personal or family history of low blood counts, bleeding diathesis, skin or nails abnormalities, unexplained liver disease, pulmonary fibrosis or alveolar proteinases, limb abnormalities, lymphedema or immune deficiency/ atypical infection.^(14)^

We were unable to confirm the presence of the suspected genetic predisposition because of lack of genetic studies but we were able to make a reasonable assumption of the possible syndromes, in eleven patients, depending on the family history and the type of familial malignancy and the presence of some non-hematological dysmorphic features. In this setting, genetic testing to rule out a familial MDS/AL predisposition syndrome is warranted to avoid use of stem cells from a relative carrying a mutation in the same MDS/AL predisposition gene ^(8,15)^.

As the current study reveals that there was no significant difference in age and gender of patient with positive family history and those with negative family history, it is recommended that all patients should be interviewed properly for positive family history of malignancies.

## Data Availability

All data produced in the present study are available upon reasonable request to the authors

